# Frailty, Mental Disorders, and Metabolic Syndrome: A Genetic Association and Mediation Mendelian Randomization Study

**DOI:** 10.1101/2024.01.16.24301316

**Authors:** Ming-Gang Deng, Kai Wang, Jia-Qi Nie, Fang Liu, Yuehui Liang, Jiewei Liu

## Abstract

**Objective:** To examine the genetic associations of metabolic syndrome (MetS) with frailty and mental disorders [depression, schizophrenia (SCZ), and bipolar disorder (BIP)], along with causality between frailty and MetS and the mediating role of mental disorders.

**Methods:** The summary-level datasets were obtained from recent genome-wide association studies. The genetic correlation was explored from the perspectives of global and local genetic correlation. Univariate Mendelian Randomization (UMR) was used to investigate the causal link between frailty and metabolic syndrome (MetS), followed by multivariate MR to address the confounding effects of body mass index (BMI) and physical activity (PA). Finally, two-step MR analyses were conducted to examine whether the causal relationship was mediated by mental disorders.

**Results:** The global genetic correlation analyses presented MetS was positively associated with frailty and depression, but reversely related to SCZ. Similarly, MetS was locally correlated to frailty, depression, and SCZ in numerous genomic regions. The UMR demonstrated that fragile people have a higher likelihood of suffering from MetS (OR: 2.773, 95% CI: 1.455-5.286, *p* = 0.002), and reversely people with MetS tended to be more fragile (beta: 0.211, 95% CI: 0.180-0.241, *p* < 0.001). This bidirectional causal association still existed even after adjusting for BMI and PA. The mediation analyses implied this causality was mediated by depression, but not SCZ and BIP.

**Conclusion:** Our research provided evidence of genetic correlations between MetS and frailty, depression, and SCZ. Additionally, we discovered a bidirectional causality between frailty and MetS, with depression playing a significant mediating role.

## 1. Introduction

Frailty, defined as a decrease in function across various physiological systems coupled with heightened sensitivity to stressors, is experiencing a significant rise due to the aging population (1). Concurrently, frailty is linked to numerous negative health outcomes and a surge in healthcare expenditures (2; 3). Metabolic syndrome (MetS), which was initially recognized as a prevalent metabolic disorder stemming from the rise in obesity rates, is now acknowledged by the International Diabetes Federation as a constellation of risk factors for cardiovascular diseases and type 2 diabetes (4). Its contribution to a range of non-communicable chronic diseases results in a substantial disease burden, underscoring the importance of early prevention.

The positive association between frailty and MetS has gained wide recognition through numerous epidemiological studies. Notably, a meta-analysis comprising 11 observational studies revealed a significant correlation between MetS and the occurrence of frailty (5). Furthermore, in a cohort study of 1,499 older adults free from diabetes at the outset, those with MetS were found to have a higher risk of developing frailty (6). Additionally, findings from the Irish Longitudinal Study on Ageing indicated an escalated risk of developing frailty over a period of four years associated with MetS (7). However, the precise causative relationship between frailty and MetS remains insufficiently explored as the observational study design was susceptible to confounding bias and reverse causality.

Recent research has shed light on the complex development of frailty and MetS, focusing on insulin resistance as a central explanatory factor. At the same time, studies have revealed a significant connection between insulin resistance and various mental health disorders, including depression(8), schizophrenia (SCZ)(9), and bipolar disorder (BIP)(10). Additionally, our previous study has shown the causal bidirectional causal association of frailty with depression and SCZ, leading to an in-depth exploration of the potential mediating role of mental health disorders in explaining the causal link between frailty and MetS. Ongoing investigations into the mediating role of mental disorders have the potential to provide valuable insights into the complex interconnections among these conditions.

Large-scale genome-wide association studies (GWASs) have identified numerous genetic loci associated with frailty, MetS, and mental disorders, which could enhance the statistical power and offer the opportunity to explore their genetic associations. In addition, Mendelian Randomization (MR), which uses genetic variants (single nucleotide polymorphisms, SNPs) as instruments, is a powerful method for obtaining robust causal inference and has been widely utilized in recent epidemiological research(11; 12). As a result, this study aimed to investigate the genetic association and causality between frailty and MetS and to check whether mental disorders played mediation roles.

## 2. Method and materials

### 2.1. Study design

An outline of the study design for this research is presented in **Fig 1**. The study was conducted in two parts: The first part involved exploring the genetic correlation of MetS with frailty and mental disorders through global and local genetic correlation analyses. The second part aimed to examine the causal relationship between frailty and MetS and determine whether mental disorders played a mediating role. The causal association between frailty and MetS was initially assessed via univariate MR (UMR) analyses, followed by multivariate MR (MVMR) analyses with adjustments for confounders. The mediating roles of mental disorders were evaluated through two-step MR analyses.

**Fig 1.**
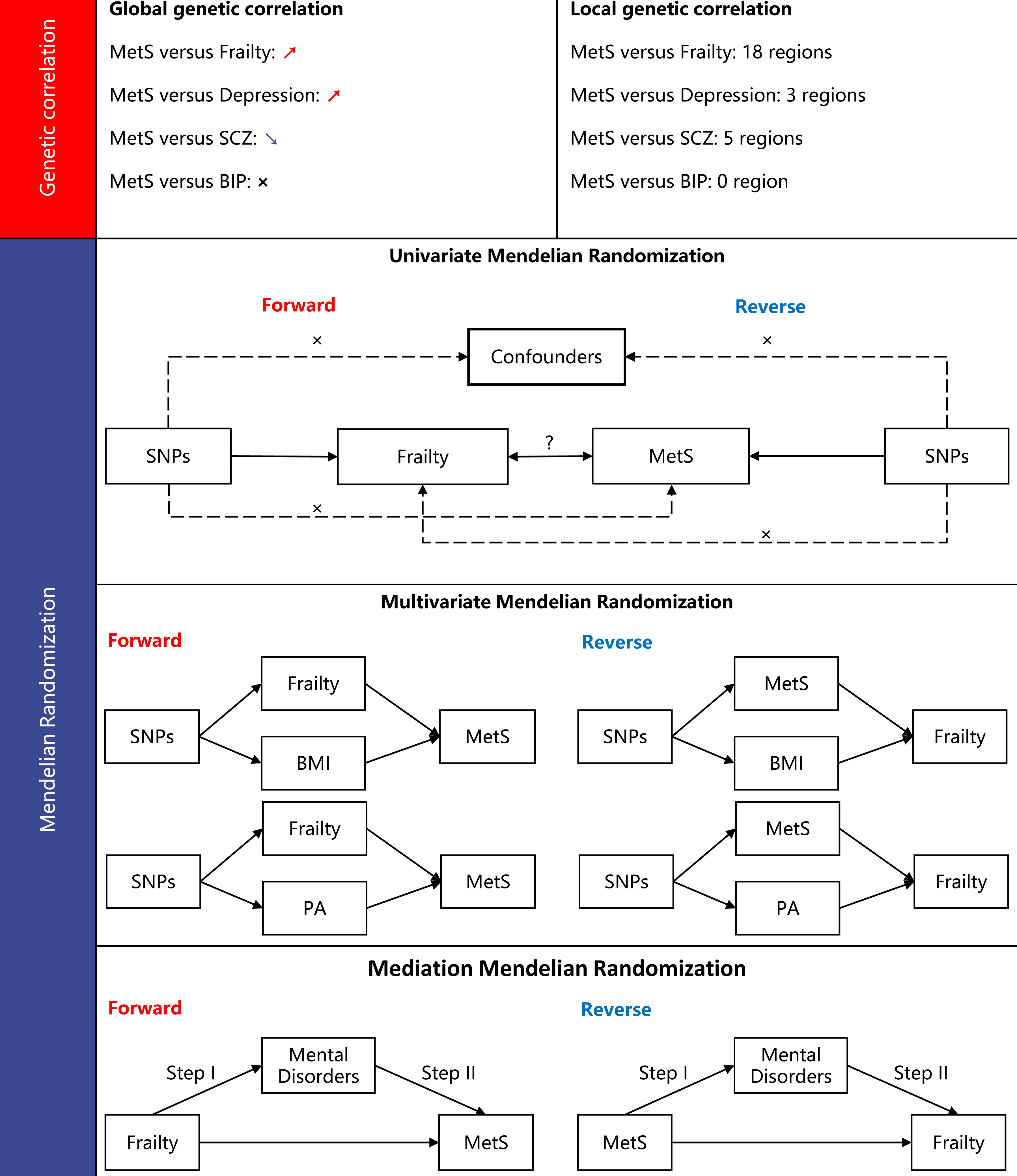
Study overview. **Abbreviations:** MetS, metabolic syndrome; SCZ, schizophrenia; BIP, bipolar disorder; SNPs, single nucleotide polymorphisms; BMI, body mass index; PA, physical activity.

### 2.2. Data sources

The genetic variants associated with frailty were derived from a recent large-scale GWAS study of fried frailty scores, as delineated by the five criteria: weight loss, exhaustion, physical activity, walking speed, and grip strength(13). The study included 386,565 participants of European descent enrolled in the UK Biobank, whose average age was 57 years old and women made up 54% of the total.

The genetic data related to MetS are based on the latest study from the Complex Trait Genetics Lab (CTG), which contains 461,920 valid subjects of European ancestry(14). The researchers systematically analyzed genetic correlations of fasting glucose, high-density lipoprotein (HDL) cholesterol, systolic blood pressure, triglycerides, and waist circumference using the structural equation modeling method, and identified 235 loci related to MetS.

For the mental disorders, we focused on depression, schizophrenia (SCZ), and bipolar disorder (BIP), which were common and have been linked to MetS. Data related to depression was obtained from the Integrative Psychiatric Research (iPSYCH) consortium, which included 166,773 cases and 507,679 controls(15). The genetic data related to SCZ and BIP were retrieved from the Psychiatric Genomics Consortium (PGC). The GWAS of SCZ was composed of 52,017 individuals with SCZ and 75,889 controls(16), while the BIP data included 40,463 people with BIP and 313,436 controls(17).

### 2.3. Genetic association

We evaluated the genetic association of MetS with frailty and mental disorders from two perspectives: global and local genetic correlation. The global genetic correlation was calculated using linkage disequilibrium score regression (LDSC), which allowed us to assess the cross-trait genetic correlation from GWAS summary statistics. The estimate (*r*_g_) varies from −1 to +1, with −1 indicating a complete negative correlation, and +1 representing a perfect positive correlation.

SUPERGNOVA (SUPER GeNetic cOVariance Analyzer), a statistical framework to perform local genetic correlation analysis, was used to quantify the genetic similarity of two traits in genomic regions. The algorithm partitions the entire genome into approximately 2,353 LD-independent blocks and offers a precise quantification of the similarity between pairs of traits influenced by genetic variations in each region. The statistical significance threshold for testing the local genetic correlation was corrected by the Bonferroni method.

### 2.4. Mendelian Randomization

#### 2.4.1. Instrumental variants selection

SNPs were Initially screened utilizing a threshold of genome-wide significance (*p* < 5×10^-8^) to identify robust relationships with exposures. The F statistic was then calculated to determine if the SNPs were susceptible to weak instrument bias, and the instruments having an F statistic of less than 10 were eliminated.

Subsequently, the PLINK v1.9 was employed to clump for linkage disequilibrium (LD) with the European samples from the 1000 Genome Project as the reference panel, and the clumping R^2^ cutoff was set to 0.001 within a window size of 10,000 kb. If the LD effect was identified, only the SNP with the lowest *p*-value would be kept.

We then harmonized the exposure and outcome datasets, where the palindromic SNPs were excluded. If any instrument was unavailable in the outcome data, it would be replaced by the proxied genetic variations (minimum LD R^2^ = 0.8 and minor allele frequency threshold = 0.3). After the aforementioned procedures were completed, the remained SNPs were utilized for conducting the MR analysis.

#### 2.4.2. Statistical analyses

We first performed the UMR to assess the causal relationship between frailty and MetS. The inverse variance weighted (IVW) method was treated as the primary method to obtain the causal estimate. This method assumes the presence of an average pleiotropic effect and is the most efficient method in this case. Prior to performing the IVW analyses, Cochran’s Q statistic was calculated to evaluate whether heterogeneity existed among the genetic variants. If significant heterogeneity was detected (*p*-value < 0.05), the causal effect would be estimated under the random-effect IVW method; otherwise, the fixed-effect method would be used. In addition to the IVW method, three pleiotropy robust methods including MR-Egger, weighted median, and MR-LASSO were employed to evaluate the stability of the results. The leave-one-out (LOO) analyses and MR-Egger regression intercept term were respectively conducted to check whether the overall effect was driven by a single SNP or to assess the possible presence of horizontal pleiotropy.

Apart from the UMR, the MVMR analyses were conducted to obtain the causal inference with adjustments for the confounding effect. MVMR as an extension of UMR, using genetic variants associated with multiple and potentially related exposures, could detect the causal effects of multiple risk factors jointly. Before performing the MVMR analyses, the PhenoScanner was employed to evaluate which confounders the selected SNPs were associated with (*p* < 1×10^-5^). Body mass index (BMI) was discovered to be related to the instrumental variants associated with frailty, as well as MetS. In addition to BMI, physical activity (PA) as a common and easily modifiable lifestyle factor has been displayed to be associated with the risks of both frailty and MetS. As a result, we estimated the causal effect between frailty and MetS with adjustments for BMI and PA, and the IVW, MR-Egger, weighted median, and MR-LASSO methods were adopted.

Following the UMR and MVMR to assess the relationship between frailty and MetS, we applied the two-step UMR analyses to explore whether the mental disorders could mediate this association. When frailty was the exposure, the first step was to obtain the causal effect of frailty on mental disorders, while the second step was to obtain the causal effect of mental disorders on MetS. As for the reverse analyses, the first and second steps were to estimate the causal effects of MetS on mental disorders, and mental disorders on frailty, respectively. Similar to the above MR analyses, the IVW Method was employed as the primary method for causal inference, and the MR-Egger, weighted median as well as MR-LASSO were supplemented. The mediating effect proportions were calculated based on the estimates from the IVW method.

The MR analyses were conducted in in R software (version 4.3.1, R Foundation for Statistical Computing) with related *TwoSampleMR* (version 0.5.8), and *MendelianRandomization* (version 0.9.0) packages. Effect estimates were reported in *β* values with 95% confidence interval (CI) when the outcome was frailty and were converted to odds ratio (OR) when the outcome was the others. All statistical tests were two-tailed, and *α* = 0.05 was considered as the significant level.

## 3. Results

### 3.1. Genetic association analyses

The results of global genetic correlation analyses by the LDSC presented that genetically predicted MetS was positively related to frailty (*r*_g_ = 0.640, se = 0.026, *p* = 2.83×10^-134^), depression (*r*_g_ = 0.194, se = 0.028, *p* = 6.45×10^-12^), and negatively related to SCZ (*r*_g_ = −0.103, se = 0.020, *p* = 1.36×10^-7^), but not correlated to BIP (*r*_g_ = −0.030, se = 0.021, *p* = 0.157).

Similar to the global genetic correlation results, the local genetic correlation analyses demonstrated that MetS was locally correlated with frailty, depression, and SCZ, while no locally correlated genomic regions were identified between MetS and BIP (**Fig 2**). In particular, MetS was correlated with frailty in 18 genomic regions after correcting for multiple tests, and the estimates were all positive. Similarly, MetS was positively associated with depression in the three significant regions, while negatively related to SCZ in the 2 of 5 significant regions.

**Fig 2.**
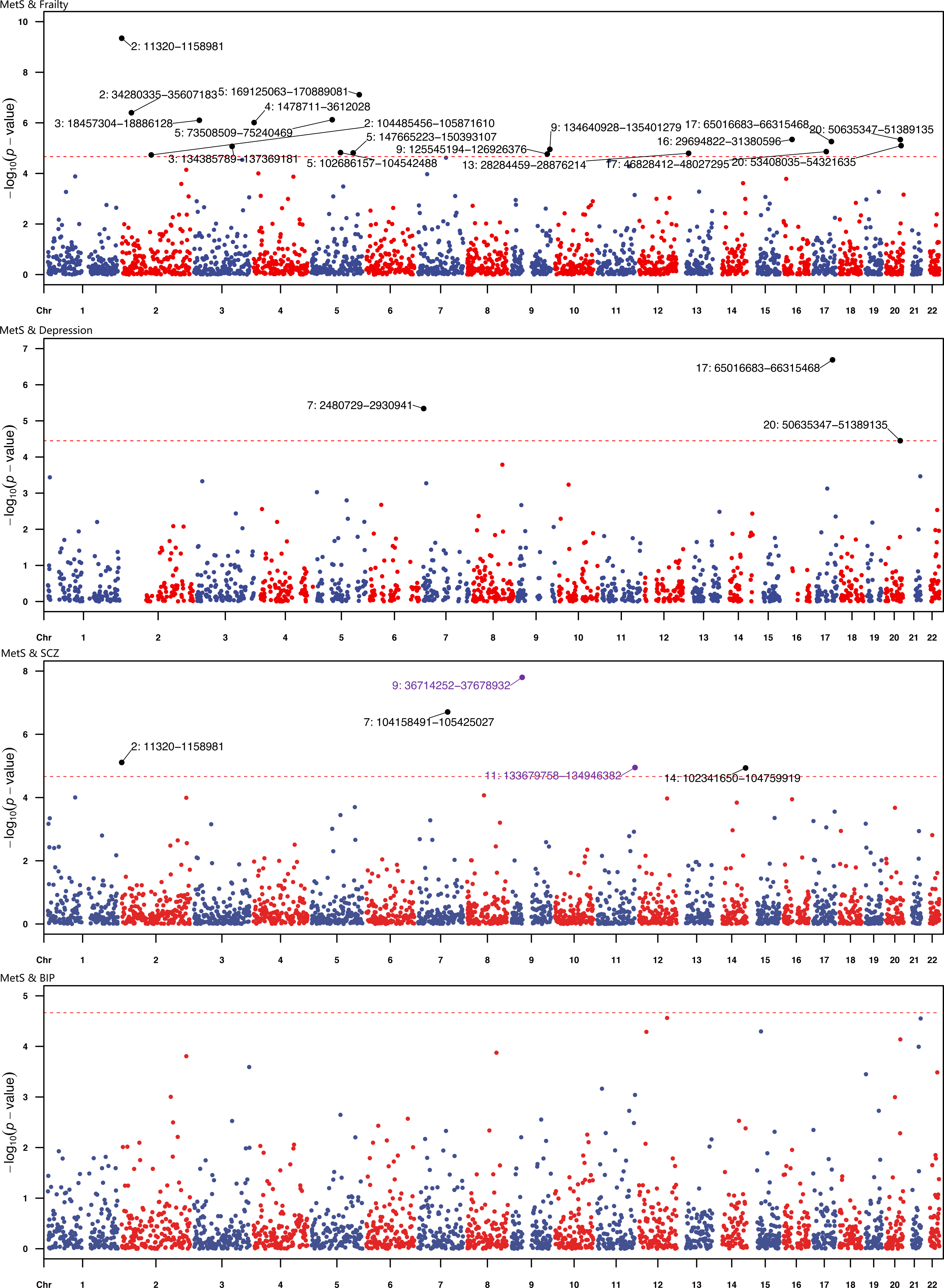
The results of local genetic correlation. The red dotted line represents the corrected significance threshold, the purple points indicate negatively correlated regions, and the black points were positively correlated regions. **Abbreviations:** MetS, metabolic syndrome; SCZ, schizophrenia; BIP, bipolar disorder

### 3.2. Mendelian Randomization analyses

#### 3.2.1. Causal relationship between frailty and MetS

The results of UMR to obtain the causality between frailty and MetS are presented in **Table 1**. The Cochran’s Q test indicated there was significant heterogeneity among the instruments related to frailty (Cochran’s Q = 581.184, *p* < 0.001) and MetS (Cochran’s Q = 649.002, *p* < 0.001). Therefore, the primary effect sizes were estimated by the random-effect IVW method. The IVW method showed that genetically predicted frailty was positively correlated with the risk of MetS (OR: 2.773, 95% CI: 1.455-5.286, *p* = 0.002), and this causality was supported by the weighted median (OR: 2.065, 95% CI: 1.692-2.519, *p* <0.001), and MR-LASSO methods (OR: 1.994, 95% CI: 1.618-2.457, *p* <0.001). The reverse analyses indicated that people with MetS were meanwhile more likely to be fragile (beta = 0.211, 95% CI: 0.180-0.241, *p* < 0.001), and the supplementary methods including MR-Egger (beta: 0.185, 95% CI: 0.100-0.270, *p* <0.001), weighted median (beta: 0.209, 95% CI: 0.173-0.244, *p* <0.001), and MR-LASSO (beta: 0.232, 95% CI: 0.214-0.251, *p* <0.001) presented similar estimates. Sensitivity analyses conducted by the MR-Egger regression intercept term presented there was no obvious horizontal pleiotropy among the SNPs related to frailty (*p* = 0.895) and MetS (*p* = 0.520), and the LOO analyses showed the bidirectional causal association between frailty and MetS were not driven by a single SNP **(Supplementary Fig S1 and Table S1)**.

**Table 1.** Univariate causal inference between frailty and metabolic syndrome.

| Exposure | Outcome | Method | beta / OR (95% CI) | *p*-value | Cochran's Q | *p*-value for heterogeneity*<br>or pleiotropy† |
| --- | --- | --- | --- | --- | --- | --- |
| Frailty | MetS | IVW | 2.773 (1.455, 5.286) | 0.002 | 581.184 | <0.001 |
|  |  | MR-Egger | 2.337 (0.179, 30.447) | 0.517 |  | 0.895 |
|  |  | Weighted Median | 2.065 (1.692, 2.519) | <0.001 |  |  |
|  |  | MR-LASSO | 1.994 (1.618, 2.457) | <0.001 |  |  |
| MetS | Frailty | IVW | 0.211 (0.18, 0.241) | <0.001 | 649.002 | <0.001 |
|  |  | MR-Egger | 0.185 (0.100, 0.270) | <0.001 |  | 0.520 |
|  |  | Weighted Median | 0.209 (0.173, 0.244) | <0.001 |  |  |
|  |  | MR-LASSO | 0.232 (0.214, 0.251) | <0.001 |  |  |

**Abbreviations:** OR, odds ratio; MetS, metabolic syndrome; IVW, inverse variance weighted.
\**p*-value for heterogeneity based on Cochran's Q statistic.
†*p*-value or pleiotropy based on MR-Egger regression intercept.

Afterward, the MVMR was conducted to determine the robustness of this bidirectional causal relationship after accounting for the confounding effects of BMI and PA. The results are detailed in **Supplementary Table S2**. After adjusting for BMI, the IVW method indicated that genetically predicted frailty was still associated with a 61.0% higher likelihood of developing MetS (OR: 1.610, 95% CI: 1.275-2.034, *p* < 0.001). Conversely, individuals with MetS were found to be more fragile (beta: 0.171, 95% CI: 0.119-0.224, *p* < 0.001). The bidirectional causal association was confirmed by the MR-Egger, weighted median, and MR-LASSO methods. Furthermore, this correlation persisted even after controlling for PA, which was corroborated by the four methods.

#### 3.2.2. Mediating role of mental disorders

By utilizing the two-step MR analyses, we assessed whether mental disorders could mediate the bidirectional causality between frailty and MetS. The results are presented in **Fig 3** and **Supplementary Table S3-S4**.

**Fig 3.**
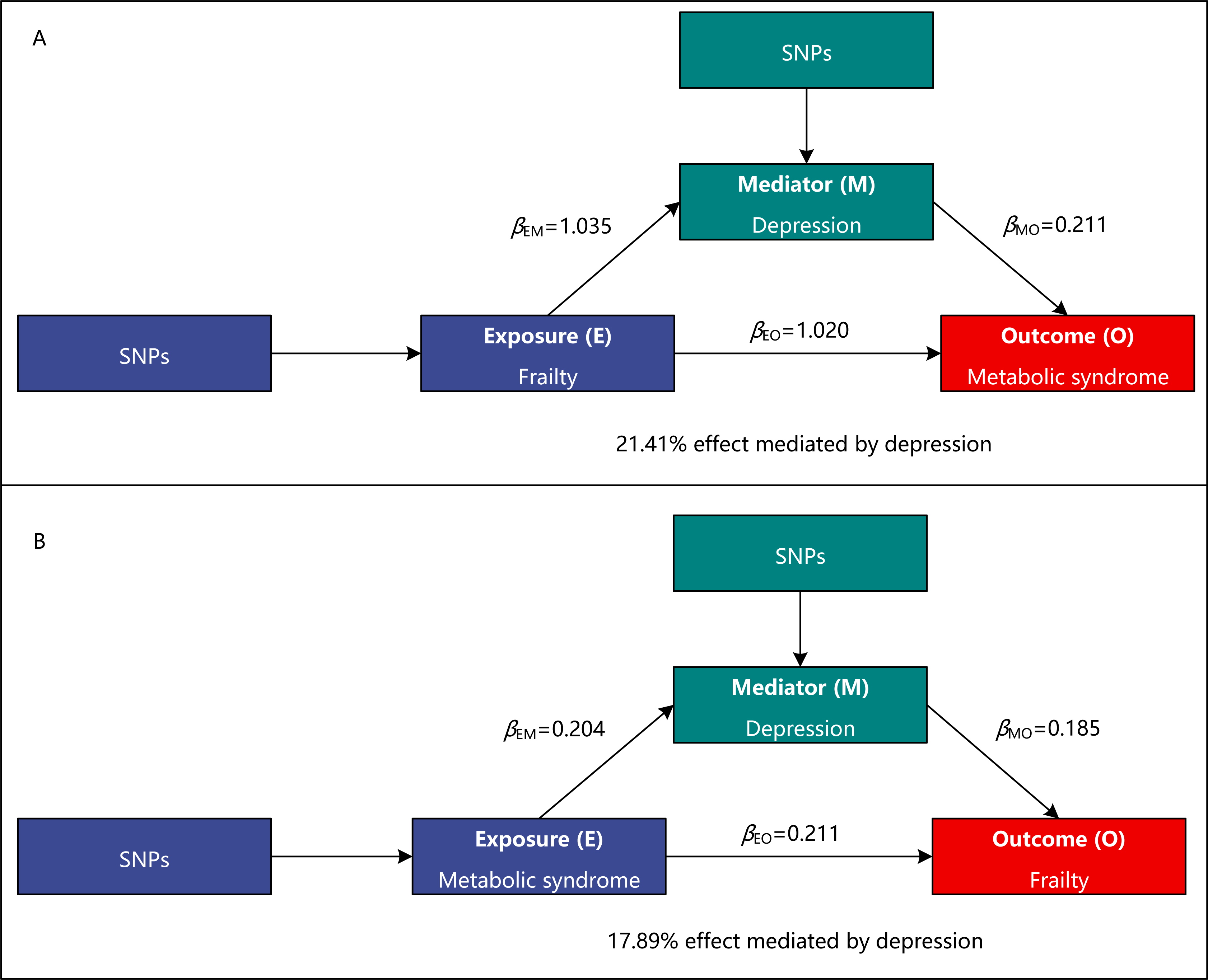
Depression mediated the bidirectional causality between frailty and metabolic syndrome.

For the forward analyses, genetically predicted frailty was positively correlated with the risk the depression (OR: 2.815, 95% CI: 1.639, 4.836, *p* <0.001), SCZ (OR: 2.230, 95% CI: 1.093, 4.549, *p* = 0.028), and depressed individuals have a higher likelihood to developing MetS (OR: 1.235, 95% CI: 1.099, 1.388, *p* <0.001). However, no other significant causal associations were discovered. Therefore, depression exerted a mediation role in the causal effect of frailty and MetS.

In the reverse analyses, it was found that individuals with MetS were discovered were more likely to suffer from depression (OR: 1.226, 95% CI: 1.117, 1.347, *p* <0.001), but not SCZ and BIP. Besides, people with depression (beta: 0.185, 95% CI: 0.144, 0.226, *p* <0.001) and SCZ (beta: 0.015, 95% CI: 0.005, 0.024, *p* = 0.002) were more likely to be frail. Similarly, the causal effect of MetS on frailty was mediated by depression.

## 4. Discussion

Our study investigated the genetic associations of MetS with frailty and mental disorders, along with the causative relationship between frailty and MetS, and the mediating effects of mental disorders. Our research furnished evidence of genetic correlations between MetS and frailty, depression, and SCZ. Furthermore, we uncovered a bidirectional causality between frailty and MetS, with depression playing a significant mediating role.

While Barzilay et al. found no association between MetS and incident frailty(18), most studies confirm a significant link(5; 19). Meanwhile, a similar study by Jiahao Zhu et al. demonstrates the bidirectional causal association of frailty with cardiometabolic diseases, including coronary artery disease, stroke, and type 2 diabetes(20). Nevertheless, MetS in the current study did not fully align with cardiometabolic diseases. Collectively, frailty and MetS exhibit associations rooted in the various components of MetS, evidenced by abdominal obesity(21), high blood pressure(22), remnant cholesterol(23), and low HDL-cholesterol(24). In addition to the above factors, the U-shape association of serum glucose level and frailty was determined in older adults without diabetes(25).

Contemporary researchers are increasingly delving into the molecular mechanisms underlying both frailty and MetS, uncovering shared pathogenic pathways. One prominently explored commonality is insulin resistance, recognized as a pivotal factor in both conditions. Emerging evidence underscores the pivotal role of insulin-like growth factor (IGF) in the hypothalamic-pituitary axis(26), contributing to frailty through reduced muscle protein synthesis, involving the Akt signaling pathway and AMP-activated protein kinase(27). Meanwhile, insulin plays a crucial role in muscle protein synthesis and maintenance while insulin resistance impaired relevant signaling pathways to reduce muscle protein synthesis and increase the breakdown of muscle tissue, leading to the development of frailty. Therefore, insulin resistance as the central feature of MetS is acknowledged as a precursor to frailty(26), prompting investigations into it as a target for prevention(28).

Vitamin D (VD), essential for bone health and calcium metabolism, is correlated with frailty by tremendous longitudinal studies and randomized controlled trials(29; 30). Besides, the VD/VD receptor axis regulate biological processes including proteolysis, mitochondrial function, cellular senescence, and adiposity(31), contributing to sarcopenic muscle atrophy and subsequent frailty. Furthermore, the relationship between VD deficiency and MetS was approved by plenty of studies in adults and children, and one of the potential mechanisms is the effect of VD on insulin secretion and sensitivity(32). VD could improve insulin resistance and VD deficiency compromises the capacity of pancreatic *β* cells to convert pro-insulin into insulin, while insulin serves as a vital factor in the prevention of MetS. Another recent plausible explanation is the gut microbiota-bile acid axis possibly through the modulation of the PPAR signaling pathway, as evidenced to mediate the relationship between plasma VD and MetS in 2966 Chinese participants with a 9-year follow-up(33). Hence, VD emerges as a potential link between frailty and MetS.

Oxidative stress and inflammation play parallel roles in frailty and MetS through various aspects and pathways. Oxidative stress is closely linked to aging while the most notable manifestation of aging is the clinical condition of frailty. Both meta-analyses and mouse models(34) provided robust evidence of oxidative stress and chronic inflammation as relevant biomarkers and precursors of frailty. Oxidative stress, characterized by excessive reactive oxygen species production, oxidized lipids to produce lipid peroxides, disrupted redox signaling and activated the apoptosis pathway. Moreover, oxidative stress affects insulin metabolic signaling and promotes cardiovascular and renal inflammation and fibrosis, resulting in damage to relevant organs and subsequent MetS(35). The NF-κB signaling pathway, the major regulator of the proinflammatory response, suppresses insulin metabolic signaling and promotes in establishment a low-grade chronic inflammatory state, leading to subsequent metabolic damage(36).

In addition to the aforementioned explanations, our investigation extended to include depression, SCZ, and BIP, aiming to scrutinize the mediating impact of these mental disorders. The results underscored a significant mediation effect, particularly notable in the case of depression. Depression emerges as a prevalent factor among individuals affected by frailty and MetS, and insulin resistance is implicated as a crucial contributor to the intricate relationship between frailty, MetS, and the manifestation of depressive symptoms. Supporting this exploration, a meta-analysis of 70 studies and over 240 million participants elucidated a noteworthy positive association between the insulin resistance index and acute depression, and this index remained unaltered even when depression symptoms were alleviated(37). Delving deeper into the molecular mechanisms, insulin resistance was identified as a key player in altering central dopaminergic signaling, particularly in the ventral tegmental area associated with reward behavior(38), contributing to the development of anhedonia and depression. Specifically, insulin resistance was found to diminish the synthesis and reuptake of dopamine through attenuated AKT signaling pathways, while the dysregulation of dopamine intensively promotes the pathophysiologic development of depression(39).

Our study had several noteworthy strengths. Firstly, we utilized genetic variants from large-scale GWASs, which enhanced the statistical power to detect genetic associations. Additionally, by using genetic variants as instruments, Mendelian randomization (MR) could provide valid causal inference, overcoming the limitations of conventional observational studies, such as confounding bias and reverse causality. Moreover, our study focused on mental disorders and highlighted the mediating role of depression in the relationship between frailty and MetS.

While our study has these advantages, it should be considered within the context of its limitations. Firstly, the participants in this research were all of European ancestry, which may limit the generalizability of our findings. Secondly, the different methods used to assess frailty may also influence the results. The frailty index (FI) was another frequently used tool to evaluate an individual’s frailty status, and the recent GWAS has discovered 14 loci related to FL(40). We did not use this index out of concern for biased causal inference, as high blood pressure, lipid disorder, and diabetes were important scoring items for FL and, at the same time, the main diagnostic criteria for MetS.

## 5. Conclusion

Our research provided evidence of genetic correlations between MetS and frailty, depression, and SCZ. Additionally, we discovered a bidirectional causality between frailty and MetS, with depression playing a significant mediating role.

## 6. Declarations

### Ethics approval

The data sets used in this research were publicly available from the published articles, and the ethics approvals were obtained in the original research.

## Author contributions

**Ming-Gang Deng:** Conceptualization, Methodology, Data Curation, Formal Analysis, and Writing - Original Draft; **Kai Wang:** Writing - Original Draft; **Jia-Qi Nie, Fang Liu, and Yuehui Liang:** Writing - Review & Editing; **Jiewei Liu:** Conceptualization, and Formal Analysis.

## Data Availability

The datasets utilized in this study are accessible on the IEU OpenGWAS project (https://gwas.mrcieu.ac.uk/), figshare (https://figshare.com/s/6683396c68807fe4e729), PGC (https://pgc.unc.edu/for-researchers/download-results/), and iPSYCH (https://ipsych.dk/en/research/downloads).

## Acknowledgments

None.

## Funding

The authors received no external funding.

## Competing interests

The authors declare that they have no competing interests

## Data and code availability

The datasets utilized in this study are accessible on the *IEU OpenGWAS project* (https://gwas.mrcieu.ac.uk/), *figshare* (https://figshare.com/s/6683396c68807fe4e729), *PGC* (https://pgc.unc.edu/for-researchers/download-results/), and *iPSYCH* (https://ipsych.dk/en/research/downloads).

Additionally, the code employed in this study is also available: *ldscr* (https://github.com/mglev1n/ldscr), *SUPERGNOVA* (https://github.com/qlu-lab/SUPERGNOVA), *TwoSampleMR* (https://mrcieu.github.io/TwoSampleMR/), and *MendelianRandomization* (https://github.com/cran/MendelianRandomization).

## Notes

### Competing Interest Statement

The authors have declared no competing interest.

### Funding Statement

This study did not receive any funding

