## Supplementary Fig S1 for "Frailty, Mental Disorders, and Metabolic Syndrome: A Genetic Association and Mediation Mendelian Randomization Study"

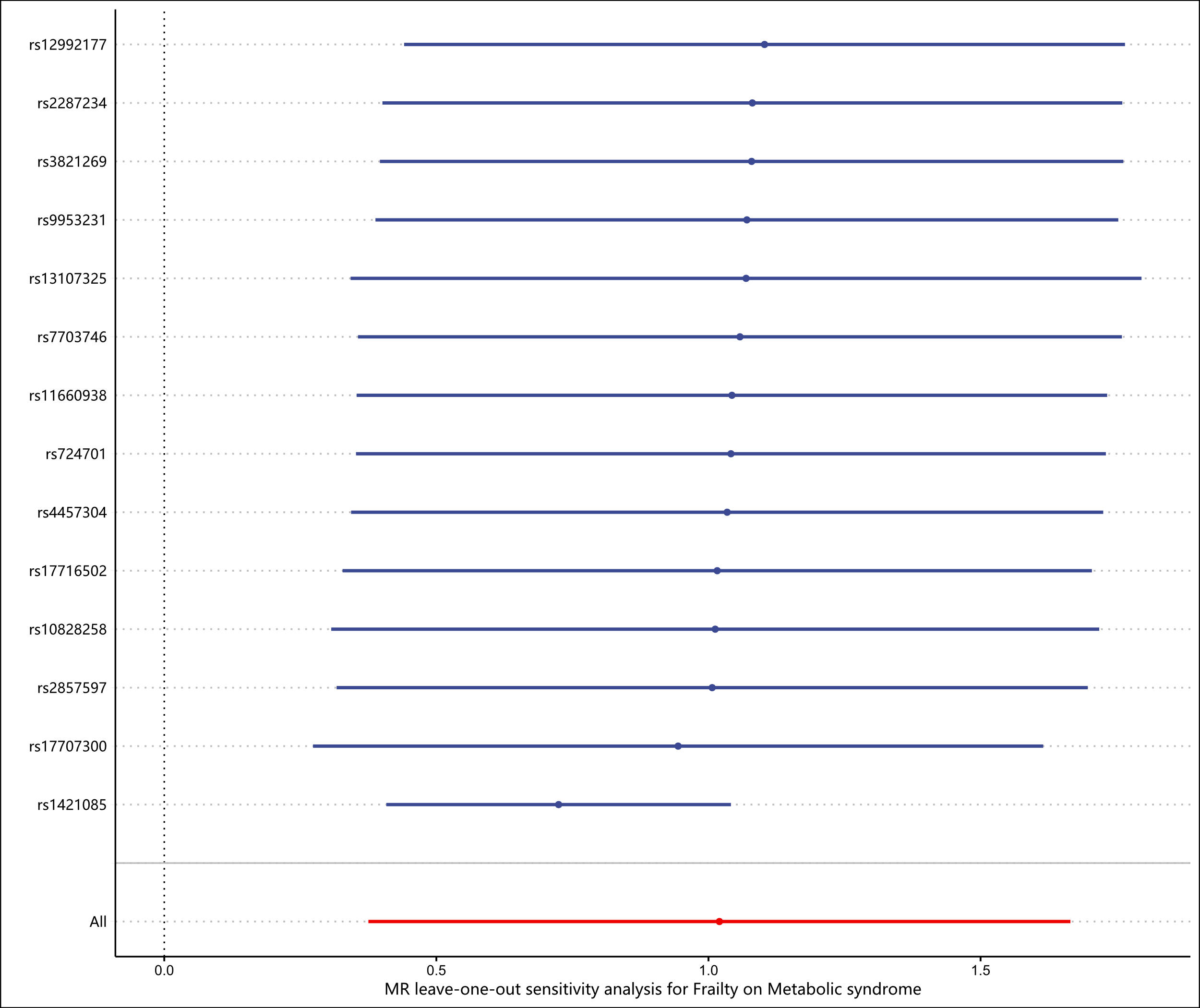

Supplementary Fig S1. Leave-one-out analysis for the causal effect of frailty on metabolic syndrome.
